# Association of cardiac biomarkers, kidney function, and mortality among adults with chronic kidney disease

**DOI:** 10.1101/2023.12.12.23299886

**Authors:** Sophie E. Claudel, Sushrut S. Waikar, Deepa M. Gopal, Ashish Verma

## Abstract

**Background and Aims:** The performance of high sensitivity troponin T (hs-cTnT), hs-cTnI, and N-terminal pro-hormone brain natriuretic peptide (NT-proBNP) in patients with chronic kidney disease (CKD) is poorly understood.

**Methods:** We included adults with CKD (eGFR<60 ml/min/1.73m^2^) in the 1999-2004 NHANES. We calculated the 99^th^ percentile of hs-cTnT, hs-cTnI (Abbott, Ortho, and Siemens assays), and NT-proBNP, measured the association between eGFR and cardiac biomarker concentration, and used Cox regression models to assess the relationship between cardiac biomarkers and CVD mortality.

**Results:** Across 1,068 adults with CKD, the mean [SD] age was 71.9[12.7] years and 61.2% were female; 78.8% had elevated NT-proBNP and 42.6% had elevated hs-cTnT based on traditional clinical reference limits. The 99^th^ percentile of hs-cTnT was 122 ng/L (95% confidence interval (CI) 101-143), hs-cTnI_Abbott_ was 69 ng/L (95% CI 38-99), and NT-proBNP was 8952 pg/mL (95% CI 7506-10,399). A 10 ml/min decrease in eGFR was associated with greater increases in hs-cTnT and NT-proBNP than hs-cTnI (hs-cTnT: 27.5% increase (β=27.5, 95% CI 28.2-43.3)), NT-proBNP 46.0% increase (β=46.0, 95% CI 36.0-56.8), hs-cTnI_Siemens_ 17.9% (β=17.9, 95% CI 9.7-26.7). Each doubling of hs-cTnT, hs-cTnI, and NT-proBNP were associated with CVD mortality (hs-cTnT HR 1.62 [95% CI 1.32-1.98], *p*<0.0001; hs-cTnI_Siemens_ HR 1.40 [95% CI 1.26-1.55], *p*<0.0001; NT-proBNP HR 1.29 [95% CI 1.19-1.41], *p*<0.0001).

**Conclusions and Relevance:** Community dwelling adults with CKD have elevated concentrations of cardiac biomarkers, above established reference ranges. Of the troponin assays, hs-cTnI concentration appears to be most stable across eGFR categories and is associated with CVD mortality.

**Clinical Perspective:** *What is new?:*

- This study investigated two important questions relevant to clinical practice. First, which cardiac biomarker (hs-cTnT or hs-cTnI) and which assay is least impacted eGFR in patients with CKD. Second, whether elevated levels of cardiac biomarkers in community-dwelling patients convey prognostic information in CKD in a more representative population.
- Community dwelling adults with CKD have elevated concentrations of cardiac biomarkers, above established reference ranges. Of the troponin assays, hs-CTnI concentration appears to be most stable across the eGFR categories and is associated with CVD mortality in individuals with CKD.

*What are the clinical implications?:*

- Community dwelling adults with CKD had elevated levels of cardiac biomarkers suggestive of subclinical myocardial injury, which were associated with increased risk of cardiovascular death.
- hs-cTnI was least affected by eGFR and retained prognostic significance, suggesting that it may be the superior assay for clinical use in CKD.

## Introduction

Patients with chronic kidney disease (CKD) are at increased risk of cardiovascular events and cardiovascular disease (CVD) mortality. High sensitivity troponins and N-terminal pro-hormone brain natriuretic peptide (NT-proBNP) are routinely used in clinical practice for diagnosis of cardiopulmonary conditions. However, patients with CKD often have elevated troponin concentrations in the absence of active coronary ischemia or other acute pathology.^1,2^ Similarly, patients with CKD often have chronically elevated NT-proBNP, which complicates the diagnosis of heart failure in this population.^3^ The prevailing assumption is that reduced biomarker clearance in the setting of low glomerular filtration rate (GFR) leads to clinically insignificant elevations in plasma concentration. However, prior work has explored variation in cardiac biomarker concentration by eGFR in CKD. Prior analyses examining cardiac biomarker concentrations in CKD are restricted to predominantly non-Hispanic White populations, have limited follow up or lack mortality outcomes, lack covariates for social determinants of health and inflammation which may confound analyses,^4,5^ and do not include any assays of high sensitivity cardiac troponin I.^6–8^

This study aims to answer two questions relevant to clinical practice. First, which cardiac biomarker (hs-cTnT or hs-cTnI) and which assay is least impacted eGFR in patients with CKD. Second, whether elevated levels of cardiac biomarkers in community-dwelling patients convey prognostic information in CKD in a more representative population.

## Methods

### Study population

This study included adults aged ≥20 years who participated in the 1999-2004 National Health and Nutrition Examination Survey (NHANES), a national, biannual, population-based study of approximately 5,000 adults each survey cycle.^9^ We excluded those who had undergone prior dialysis or were pregnant. We excluded patients without CKD based on an estimated glomerular filtration rate (eGFR) ≥60ml/min/1.73m^2^ using the 2021 CKD-EPI creatinine equation.^10^ The final sample size was N=1,068. For additional analysis comparing eGFR estimating equations, we also calculated the eGFR using the 2021 CKD-EPI creatinine-cystatin c equation^10^ and the 2012 CKD-EPI cystatin c equation.^11^ These equations were also used to restrict the population to those with eGFR <60 ml/min per 1.73m^2^ for analysis, resulting in N=939 using eGFR_cr-cys_ and N=1,231 using eGFR_cys_. This study followed the Strengthening the Reporting of Observational Studies in Epidemiology (STROBE) reporting guidelines.

### Cardiac biomarkers

High-sensitivity troponin T (hs-cTnT), high-sensitivity troponin I (hs-cTnI), and NT-proBNP were measured in a laboratory at the University of Maryland School of Medicine. We selected only the samples which had not undergone a prior freeze-thaw cycle (excluded N=19 samples).

Hs-cTnT was measured using a Roche Cobas e601 (Elecsys reagents), which has a lower limit of detection (LLOD) of 3 ng/L. hs-cTnI_Abbott_ was measured using an Abbott ARCHITECT i2000SR and had a LLOD of 1.7 ng/L. hs-cTnI_Ortho_ was measured using an Ortho Vitros 36000 and had a LLOD of 0.39 ng/L. hs-cTnI_Siemens_ was measured using a Siemens Centaur XP and had a lower limit of detection of 1.6 ng/L. NT-proBNP was measured using a Roche Cobas e601 autoanalyzer (LLOD 5 pg/ml, upper LOD 35,000 pg/ml). Details including coefficients of variability for each assay are publicly available at https://wwwn.cdc.gov/Nchs/Nhanes/1999-2000/SSTROP_A.htm.

### Outcome variables

We first examined cardiac biomarker concentration across eGFR categories and clinical variables. For longitudinal analyses, cardiovascular and all-cause mortality were determined through linkage with the National Center for Health Statistics National Death Index through December 31^st^, 2019. Cardiovascular mortality included death from heart disease (ICD-10 codes I00-I09, I11, I13, I20-I151) and cerebrovascular diseases (ICD-10 codes I60-I69).

### Other variables of interest

The NHANES survey includes additional demographic and lifestyle variables, which we included based on clinical and biological relevance. Prevalent CVD was defined as a self-reported history of angina, myocardial infarction, heart failure, stroke, or heart disease. Diabetes was defined as hemoglobin A1c >6.5% or current use of hypoglycemic medications. Three measurements of systolic and diastolic blood pressures were taken for each participant (mmHg). In this study, the systolic and diastolic blood pressures were separately averaged for each participant. Total cholesterol was measured using an enzymatic assay at Johns Hopkins University. Race or ethnicity groups provided by NHANES investigators included non-Hispanic White, non-Hispanic Black, Mexican American, or other. Smoking status was determined based on responses to several questions regarding current, former, and lifetime smoking. Food insecurity was determined by NHANES investigators based on responses to the U.S. Department of Agriculture Food Adequacy Indicator, the U.S. Household Food Security Survey Module, and participation in Special Supplemental Nutrition Programs for Women, Infants and Children. Current medications were self-reported by participants and verified by staff via prescription bottle review, when available. NHANES investigators subsequently coded individual medications into medication classes, which were used for analysis (e.g., angiotensin-receptor blocker). We included c-reactive protein (CRP) as a proxy marker for inflammation.

### Imputation

We used hot deck multiple imputation (with single random sampling without replacement) to account for complex survey design.^12^ We imputed missing values in systolic blood pressure (N=67), diastolic blood pressure (N=67), UACR (N=63), food security (N=37), BMI (N=73), eGFR (N=20), and hemoglobin A1c (N=1).

### Statistical analysis

We calculated the weighted proportion of participants with subclinical CVD based on definitions in the existing cardiovascular literature (NT-proBNP ≥125 pg/mL; hs-cTnT ≥14 ng/L in women or ≥22 ng/L in men).^13^ Subsequently, we assessed the proportion of participants with subclinical CVD by clinical eGFR category.

To assess the relationship between kidney function and cardiac biomarker concentrations, we first calculated the 99^th^ percentile of each biomarker and included bootstrapped confidence intervals (2000 replicates). The 99^th^ percentile is considered the upper limit of normal for these biomarkers in non-CKD populations.^14,15^ We examined the 99^th^ percentile in the entire CKD study population, as well as by sex, CVD history, and UACR. We then calculated the 99^th^ percentile by eGFR category (<30, 30 to <45, and 45 to <60 ml/min/1.73m^2^) within sex, race (Black vs. non-Black), and age (<60 or ≥60 years) categories.

We then modeled the association between eGFR and each biomarker individually using three separate equations to estimate GFR. The three equations used included the 2021 CKD- EPI creatinine (eGFR_cr_), 2021 CKD-EPI creatinine-cystatin c (eGFR_cr-cys_), and 2012 CKD-EPI cystatin c (eGFR_cys_) equations. Kidney function was represented as a per 10 ml/min per 1.73m^2^ reduction in eGFR. We then evaluated the relationship between eGFR and the difference between hs-cTnT and hs-cTnI (all three available assays). All models were first adjusted for age, sex, race or ethnicity, smoking, health insurance, body mass index, prevalent CVD, diabetes, hemoglobin A1c, total cholesterol, systolic blood pressure, statin use, and ACEi/ARB use, then additionally adjusted for UACR.

Finally, we used Cox proportional hazards models to assess the relationship between hs-cTnT, hs-cTnI (using all three available assays), and NT-proBNP, and mortality (cardiovascular, non-cardiovascular, and all-cause mortality). Based on clinical and biological plausibility, Model 1 was adjusted for age, sex, race or ethnicity, smoking, health insurance, body mass index, survey year, prevalent CVD, diabetes, hemoglobin A1c, total cholesterol, systolic blood pressure, statin use, and ACEi/ARB use. Model 2 was adjusted for Model 1 + UACR. Model 3 was adjusted for Model 2 + eGFR. We then adjusted for CRP as a marker of generalized inflammation. The examined non-linear relationships between eGFR and cardiac biomarkers using restricted cubic splines using ‘rms’ package in R software.^16^ We performed a sensitivity analysis assessing the associations with both cardiovascular and all-cause mortality stratified by self-reported history of CVD.

To assess whether these three cardiac biomarkers offer independent prognostic information, we adjusted models of hs-cTnT with hs-cTnI (all three available assays). Similarly, we adjusted models of hs-cTnI and NT-proBNP with hs-cTnT. We then calculated the c-statistic for the models before and after adding the additional biomarker to the model. We used Pearson correlations to evaluate correlations between the hs-cTnT and hs-cTnI assays.

Analyses were weighted using complex survey procedures and conducted using SAS software (SAS Institute, version 9.4) and R software, version 3.5 (R Project for Statistical Computing). A two-sided *p*-value <0.05 was considered statistically significant.

## Results

The mean age was 71.9 years (SD 12.7), men represented 38.8% and 76.9% were non- Hispanic White (**Table 1**). The mean eGFR was 47.3 ml/min (SD 11.0) and median UACR was 12.1 mg/g [IQR 5.9, 45.9]. Prevalence of subclinical CVD using conventional thresholds^13^ was 42.6% based on NT-proBNP and 79.1% based on sex-specific troponin thresholds in the overall population. Higher proportions were seen in the lowest eGFR categories (**Supplemental Figure 1**).

**Table 1.** Baseline characteristics of the study population.

| **Table 1. Baseline characteristics of the study population** |  |
| --- | --- |
|  | **Mean (SD), median [IQR], or weighted %** |
| Age, years | 71.9 (12.7) |
| Male sex | 38.8 |
| Race or ethnicity |  |
| Non-Hispanic White | 76.9 |
| Mexican American | 1.7 |
| Non-Hispanic Black | 13.8 |
| Other | 7.6 |
| Education |  |
| Below High School | 35.2 |
| High School/GED | 27.8 |
| Above High School | 37.0 |
| Current or former smoking | 49.8 |
| Uninsured | 3.0 |
| Food insecure | 7.4 |
| Insufficient physical activity | 73.2 |
| Diabetes | 22.8 |
| Hyperlipidemia | 52.9 |
| Hypertension | 85.0 |
| Cardiovascular disease <sup>a</sup> | 38.3 |
| eGFR ml/min per 1.73m<sup>2</sup> |  |
| 2021 CKD-EPI<sub>cr</sub> | 47.3 (11.0) |
| 2021 CKD-EPI<sub>cr-cys</sub> | 45.7 (11.8) |
| 2012 CKD-EPI<sub>cys</sub> | 45.1 (11.7) |
| UACR, mg/g | 12.1 [5.9, 45.9] |
| CKD stage based on eGFR<sub>cr</sub> |  |
| G1 (eGFR ≥90 ml/min/1.73m<sup>2</sup>) | 0 |
| G2 (eGFR 60-90 ml/min/1.73m<sup>2</sup>) | 0 |
| G3a (eGFR 45-60 ml/min/1.73m<sup>2</sup>) | 67.4 |
| G3b (eGFR 30-45 ml/min/1.73m<sup>2</sup>) | 25.0 |
| G4 and G5 (eGFR <30 ml/min/1.73m<sup>2</sup>) | 7.5 |
| Albuminuria grade |  |
| A1 (UACR <30 mg/g) | 68.6 |
| A2 (UACR 30-300 mg/g) | 21.8 |
| A3 (UACR ≥300 mg/g) | 9.6 |
| Body mass index, kg/m<sup>2</sup> | 28.5 (6.5) |
| Hemoglobin A1c, % | 5.9 (1.1) |
| Low density lipoprotein, mg/dL | 118.7 (27.7) |
| Total cholesterol, mg/dL | 205.7 (46.0) |
| Systolic blood pressure, mmHg | 140.0 (24.8) |
| Diastolic blood pressure, mmHg | 67.6 (14.7) |
| Cardiac biomarker concentrations |  |
| hs-cTnT, ng/L | 14.7 [9.6, 23.9] |
| hs-cTnI<sub>Abbott</sub>, ng/L | 4.8 [2.7, 8.5] |
| hs-cTnI<sub>Ortho</sub>, ng/L | 2.2 [1.0, 5.2] |
| hs-cTnI<sub>Siemens</sub>, ng/L | 6.8 [3.6, 12.8] |
| NT-proBNP, pg/mL | 277.2 [115.8, 672.2] |
| Subclinical CVD by conventional thresholds |  |
| % with NT-proBNP ≥125 pg/mL | 78.8 |
| % with hs-cTnT ≥14 ng/L (women) or ≥22 ng/L (men) | 42.6 |
| C-reactive protein, mg/dL | 0.3 [0.1, 0.7] |
| ACEi/ARB use | 35.8 |
| Statin use | 28.6 |
| <sup>a</sup> cardiovascular disease includes myocardial infarction, angina, heart failure, heart disease, or stroke.<br>Abbreviations: ACEi, angiotensin converting enzyme inhibitor; ARB, angiotensin receptor blocker; CKD, chronic kidney disease; eGFR, estimated glomerular filtration rate; GED, general education diploma; hs-cTnT, high-sensitivity troponin T; NT-proBNP, N-terminal b-type natriuretic peptide; UACR, urinary albumin-to-creatinine ratio. |  |

### Effect of kidney function on cardiac biomarker concentration

The 99^th^ percentile of each biomarker among those with CKD were hs-cTnT 122 ng/L (95% confidence interval (CI) 101-143 ng/L), hs-cTnI_Abbott_ 69 ng/L (95% CI 38-99 ng/L), hs-cTnI_Ortho_ 56 ng/L (95% CI 27-84 ng/L), hs-cTnI_Siemens_ 172 ng/L (95% CI 116-228 ng/L), and NT-proBNP 8,952 pg/ml (95% CI 7,506-10,399 pg/ml) (**Table 2**). Compared to hs-cTnI, values of hs-cTnT and NT-proBNP were higher at lower levels of eGFR, higher grades of albuminuria, and among those with prior CVD (**Table 2**). hs-cTnT was modestly correlated to all hs-cTnI assays, but the correlations were stronger between hs-cTnI assays (*p*<0.0001 for all; Supplemental Table 1).

**Table 2.** 99^th^ percentile of cardiac biomarkers by clinical characteristics.

|  |  |  |  |  |  |
| --- | --- | --- | --- | --- | --- |
|  | **hs-cTnT** | **hs-cTnI<sub>Abbott</sub>** | **hs-cTnI<sub>Ortho</sub>** | **hs-cTnI<sub>Siemens</sub>** | **NT-proBNP** |
| Previously proposed upper reference limit in CKD <sup>a b</sup> | 48 ng/L (24-72) <sup>a</sup><br>126 ng/L <sup>b</sup> | 42 ng/L (34-51) <sup>a d</sup> |  |  | 1,039 pg/mL <sup>a</sup><br>3,592 pg/mL <sup>b</sup> |
| Manufacturer upper reference limit | 14 ng/L (women) <sup>c</sup><br>22 ng/L (men) <sup>c</sup> | 17 ng/L (women) <sup>c</sup><br>35 ng/L (men) <sup>c</sup> | 9 ng/L (women) <sup>c</sup><br>12 ng/L (men) <sup>c</sup> | 40 ng/L (women) <sup>c</sup><br>58 ng/L (men) <sup>c</sup> | 125 pg/mL <sup>b</sup> |
| **Overall** | 122 (101, 143) | 69 (38, 99) | 56 (27, 84) | 172 (116, 228) | 8952 (7506, 10399) |
| **Overall by sex** |  |  |  |  |  |
| Men | 158 (127, 188) | 82 (0, 234) | 68 (0, 169) | 246 (71, 422) | 13742 (11758, 15727) |
| Women | 82 (62, 102) | 62 (21, 102) | 51 (6, 97) | 169 (95, 244) | 6328 (2512, 10143) |
| **Overall by CVD history** |  |  |  |  |  |
| No CVD | 85 (60, 109) | 69 (0, 199) | 57 (0, 147) | 126 (0, 269) | 5106 (1996, 8215) |
| Prior CVD | 197 (156, 239) | 71 (13, 129) | 53 (0, 109) | 197 (89, 305) | 9559 (5794, 13325) |
| **Overall by UACR** |  |  |  |  |  |
| <30 mg/g | 71 (40, 101) | 58 (0, 135) | 51 (0, 111) | 164 (67, 260) | 4081 (2185, 5976) |
| 30-300 mg/g | 102 (60, 145) | 99 (0, 276) | 66 (0, 199) | 340 (172, 507) | 12351 (9602, 15098) |
| ≥300 mg/g | 283 (249, 317) | 74 (48, 99) | 58 (38, 78) | 155 (130, 181) | 12918 (8627, 17209) |
| **Overall by eGFR<sub>cr</sub>** |  |  |  |  |  |
| 45-60 ml/min/1.73m<sup>2</sup> | 76 (43, 110) | 74 (0, 185) | 43 (0, 131) | 208 (82, 334) | 4032 (3490, 4574) |
| 30-44 ml/min/1.73m<sup>2</sup> | 111 (99, 123) | 72 (55, 89) | 69 (53, 85) | 138 (112, 164) | 12674 (10422, 14926) |
| <30 ml/min/1.73m<sup>2</sup> | 288 (245, 331) | 61 (33, 89) | 42 (36, 48) | 299 (194, 403) | 21810 (17696, 25926) |
| **Men** |  |  |  |  |  |
| 45-60 ml/min/1.73m<sup>2</sup> | 98 (59, 137) | 95 (0, 253) | 53 (0, 172) | 286 (92, 481) | 3787 (2743, 4831) |
| 30-44 ml/min/1.73m<sup>2</sup> | 118 (111, 125) | 72 (61, 83) | 70 (52, 89) | 121 (108, 133) | 15847 (12867, 18825) |
| <30 ml/min/1.73m<sup>2</sup> | 315 (252, 376) | 52 (48, 56) | 40 (37, 43) | 61 (57, 65) | 33070 (28937, 27202) |
| **Women** |  |  |  |  |  |
| 45-60 ml/min/1.73m<sup>2</sup> | 44 (41, 46) | 56 (3, 108) | 34 (0, 95) | 170 (58, 282) | 3799 (3289, 4309) |
| 30-44 ml/min/1.73m<sup>2</sup> | 93 (74, 111) | 65 (43, 86) | 54 (27, 82) | 129 (103, 156) | 7458 (5459, 9457) |
| <30 ml/min | 236 (195, 278) | 64 (32, 96) | 49 (40, 58) | 391 (286, 496) | 12224 (7480, 16967) |
| **Age <60 years** |  |  |  |  |  |
| 45-60 ml/min/1.73m<sup>2</sup> | 38 (5, 71) | 45 (40, 51) | 34 (29, 39) | 131 (103, 160) | 801 (711, 891) |
| 30-44 ml/min/1.73m<sup>2</sup> | 99 (78, 120) | 60 (47, 74) | 63 (45, 79) | 111 (83, 139) | 14036 (10310, 17762) |
| <30 ml/min/1.73m<sup>2</sup> | 228 (188, 269) | 51 (42, 60) | 40 (32, 47) | 437 (325, 549) | 12905 (9895, 15915) |
| **Age ≥60 years** |  |  |  |  |  |
| 45-60 ml/min/1.73m<sup>2</sup> | 81 (42, 119) | 80 (0, 199) | 54 (0, 148) | 243 (118, 369) | 4416 (3878, 4953) |
| 30-44 ml/min/1.73m<sup>2</sup> | 110 (97, 122) | 74 (56, 91) | 59 (42, 77) | 144 (118, 169) | 9219 (6565, 11872) |
| <30 ml/min/1.73m<sup>2</sup> | 289 (238, 341) | 63 (31, 95) | 48 (42, 54) | 54 (36, 73) | 32656 (28906, 36407) |
| **Black (Non-Hispanic)** |  |  |  |  |  |
| 45-60 ml/min/1.73m<sup>2</sup> | 67 (0, 135) | 178 (0, 380) | 124 (0, 279) | 408 (181, 635) | 3640 (3253, 4027) |
| 30-44 ml/min/1.73m<sup>2</sup> | 112 (99, 125) | 112 (88, 136) | 97 (66, 128) | 163 (121, 206) | 4701 (4115, 5305) |
| <30 ml/min/1.73m<sup>2</sup> | 331 (252, 410) | 65 (60, 69) | 58 (51, 65) | 438 (346, 531) | 28510 (22825, 34195) |
| **Non-Black** |  |  |  |  |  |
| 45-60 ml/min/1.73m<sup>2</sup> | 79 (43, 115) | 56 (0, 145) | 35 (0, 102) | 125 (12, 239) | 4025 (3450, 4600) |
| 30-44 ml/min/1.73m<sup>2</sup> | 108 (94, 121) | 67 (57, 77) | 58 (33, 79) | 123 (107, 139) | 14610 (12267, 16954) |
| <30 ml/min/1.73m<sup>2</sup> | 286 (241, 330) | 37 (0, 73) | 28 (25, 31) | 38 (15, 61) | 19754 (15333, 24,174) |
| <sup>a</sup> Kampmann, et al. 2022. *Diagnostics*.<br><sup>b</sup> Bansal, et al. 2021. *AJKD*.<br><sup>c</sup> McEvoy, et al. 2023. *Eur Heart J*.<br><sup>d</sup> Meta-analysis included multiple assays and provided a single, pooled estimate for hs-cTnI.<br>Abbreviations: ACEi, angiotensin converting enzyme inhibitor; ARB, angiotensin receptor blocker; CI, confidence interval; CVD, cardiovascular disease; eGFR, estimated glomerular filtration rate; hs-cTnI, high-sensitivity troponin I; hs-cTnT, high-sensitivity troponin T; NT-proBNP, N-terminal b-type natriuretic peptide; UACR, urinary albumin-to-creatinine ratio. |  |  |  |  |  |

The associations between cardiac biomarkers and kidney function were variable between biomarkers and eGFR estimating equations (**Figure 1**). In the multivariable adjusted model, for every 10 ml/min per 1.73m^2^ decrease in eGFR, across three estimating equations, there was a 24.5 to 27.5% increase in hs-cTnT (*p*<0.0001 for all) and a 39.1 to 46.0% increase in NT-proBNP (*p*<0.0001 for all). eGFR was less strongly associated with hs-cTnI than hs-cTnT or NT-proBNP, regardless of assay or eGFR estimating equation. This is visually demonstrated using restricted cubic splines (**Figure 2**).

**Figure 1.**
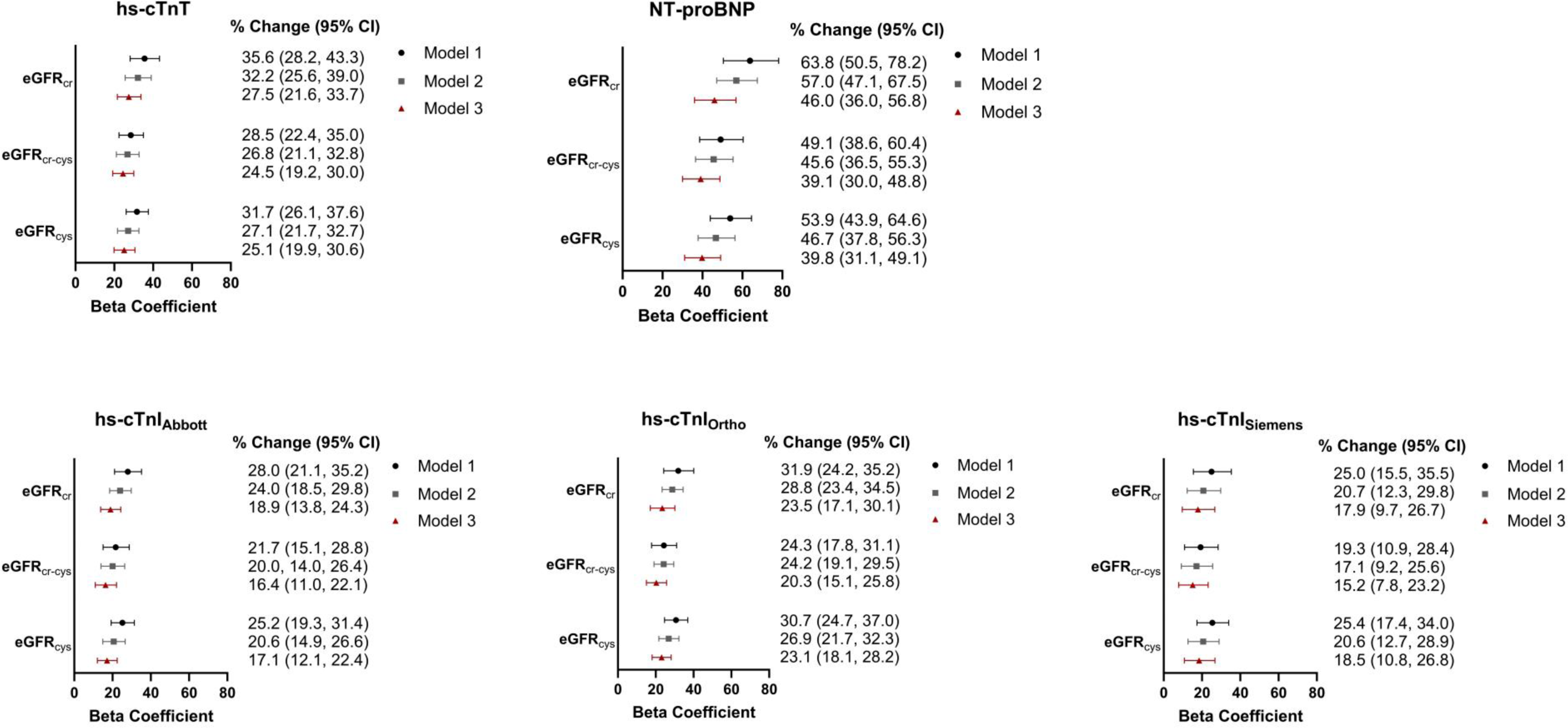
Association between eGFR and cardiac biomarker concentration, by eGFR estimating equation. Model 1 is unadjusted. Model 2 is adjusted for age, sex, race or ethnicity, insurance, smoking, history of CVD, BMI, diabetes, hemoglobin A1c, total cholesterol, systolic blood pressure, use of ACEi/ARB, use of statin. Model 3 is adjusted Model 2 + UACR. Estimating equations: eGFR_cr_ (N=1,068; 2021 CKD-EPI creatinine), eGFR_cr-cys_ (N= 939; 2021 CKD-EPI creatinine-cystatin c), eGFR_cys_ (N= 1,231; 2012 CKD-EPI cystatin c.

**Figure 2.**
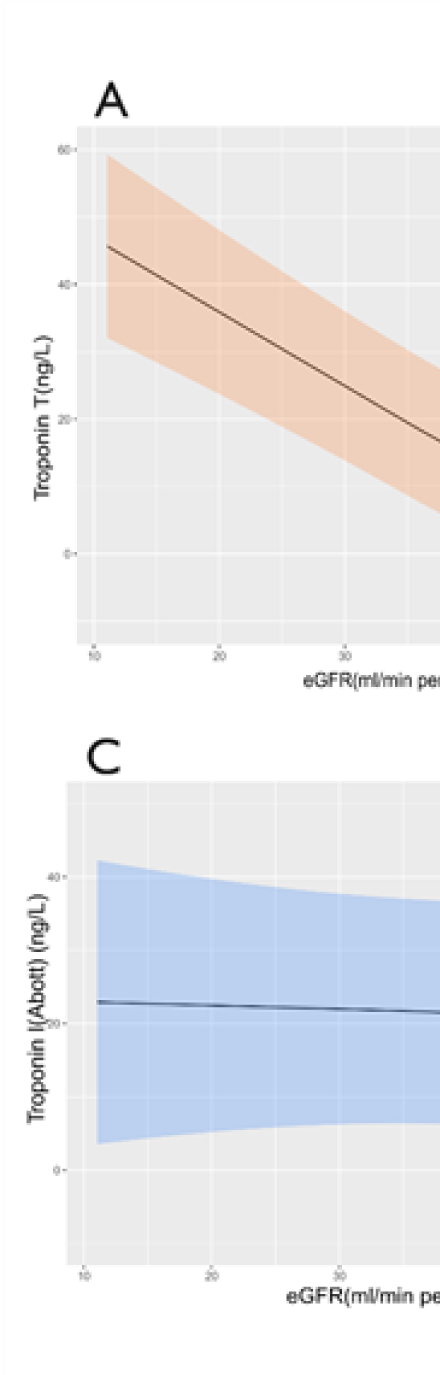
Restricted cubic splines of the relationship between eGFR and cardiac biomarkers. A, hs-cTnT; B, NT-proBNP; C, hs-cTnI_Abbott_; D, hs-cTnI_Ortho_; E, hs-cTnI_Siemens_. Restricted cubic spline model was adjusted for age, sex, race or ethnicity, insurance, smoking, history of CVD, BMI, diabetes, hemoglobin A1c, total cholesterol, systolic blood pressure, use of ACEi/ARB, use of statin, and UACR. Splines represent non-weighted data.

Examining the association between eGFR estimating equations and the difference between hs-cTnT and hs-cTnI showed differing trends based on which hs-cTnI assay was used (**Table 3**). eGFR was significantly associated with the hs-cTnT-hs-cTnI difference only when using the Abbott and Ortho assays, but not the Siemens assay. Larger differences between hs-cTnT and hs-cTnI were observed at lower eGFR (**Figure 3**).

**Figure 3.**
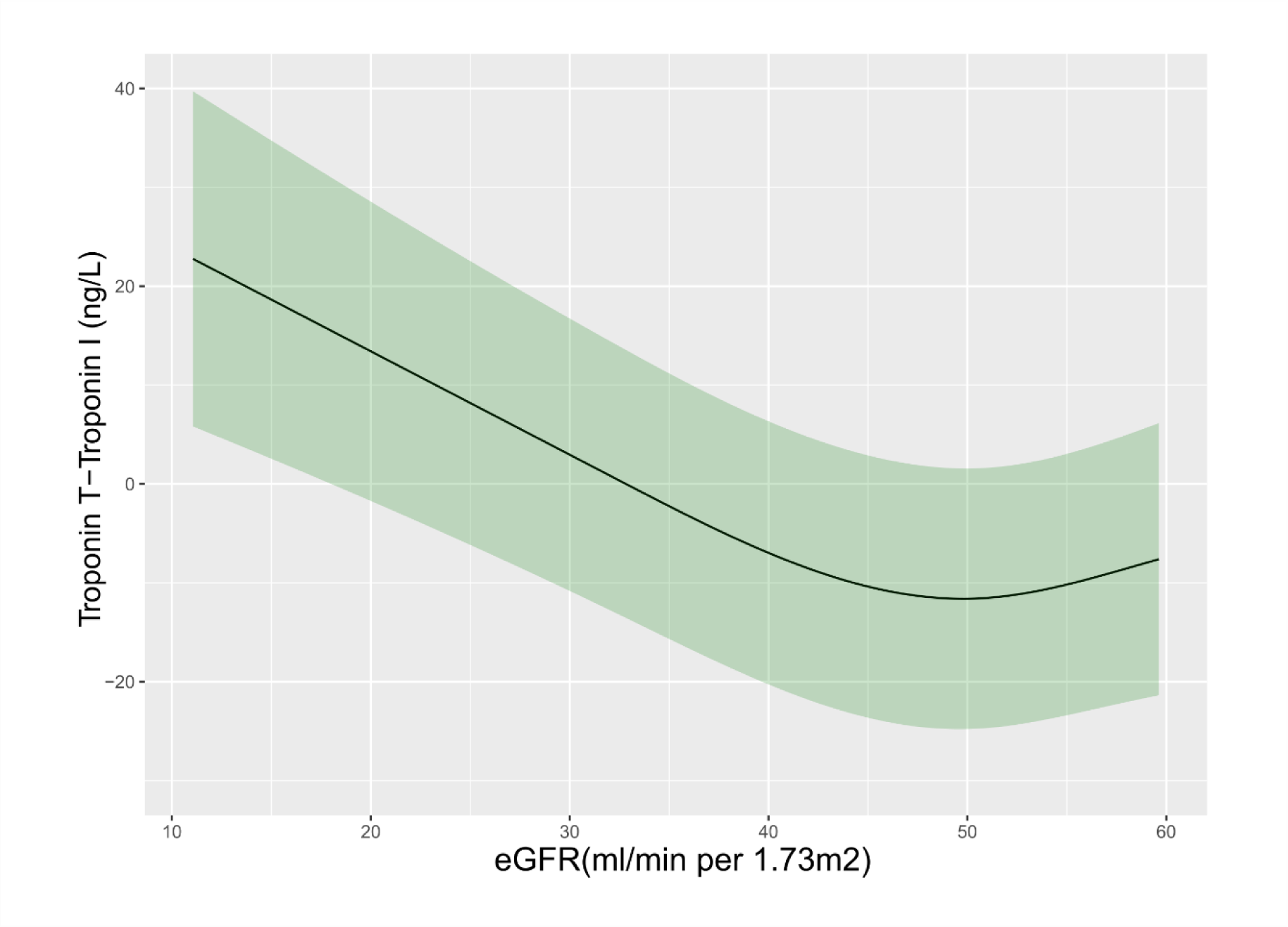
Restricted cubic splines of the relationship between eGFR and the difference between hs-cTnT and hs-cTnI_Abbott_. Restricted cubic spline model was adjusted for age, sex, race or ethnicity, insurance, smoking, history of CVD, BMI, diabetes, hemoglobin A1c, total cholesterol, systolic blood pressure, use of ACEi/ARB, use of statin, and UACR. Splines represent non-weighted data.

**Table 3.**
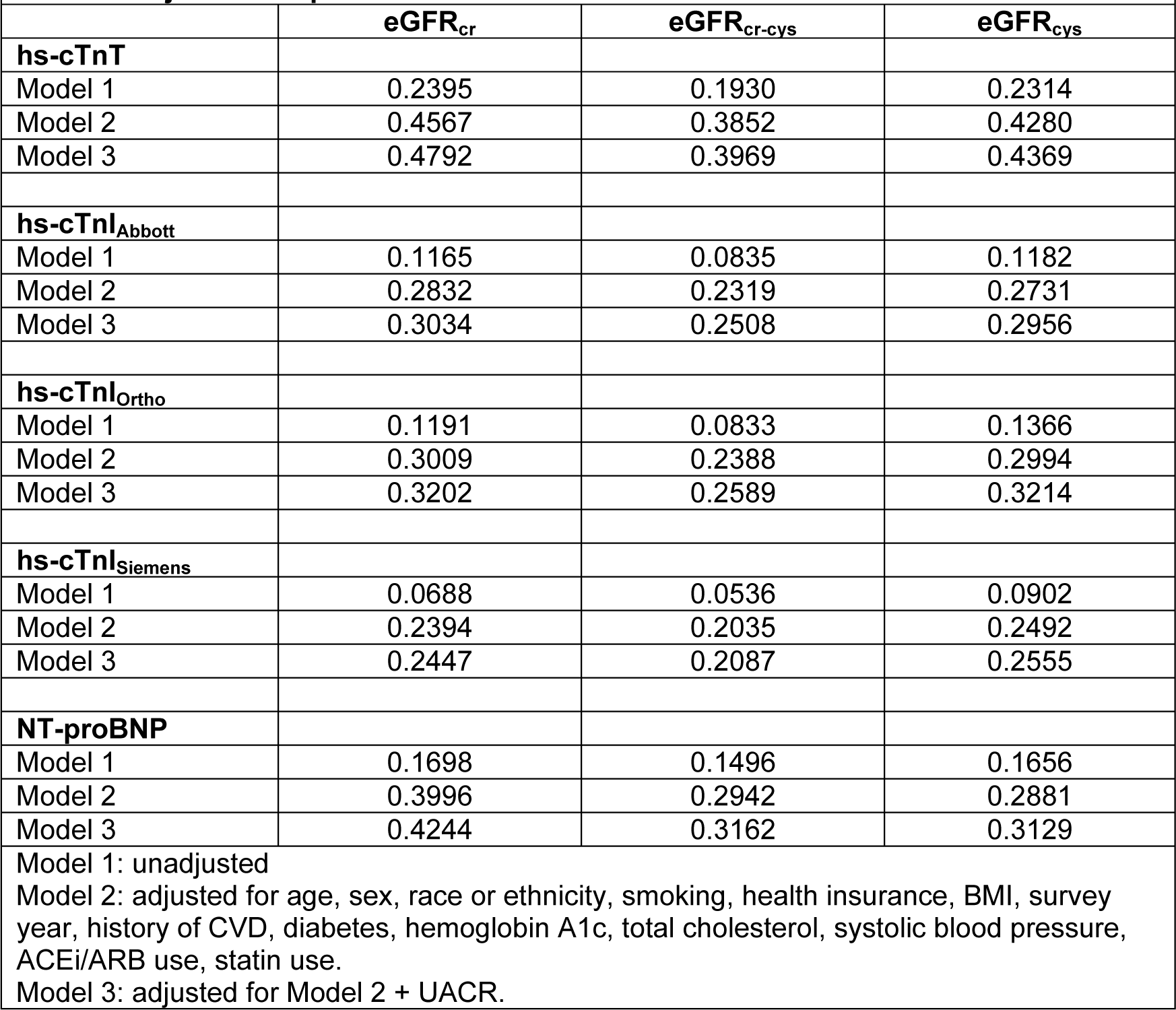
Adjusted R squared for association between eGFR and cardiac biomarkers.

### Association between cardiac biomarkers and mortality

The median follow-up time was 15.9 years and there were 864 total deaths and 336 CVD deaths. Each doubling of hs-cTnT was associated with a 62% increased risk of CVD mortality (HR 1.62 [95% confidence interval 1.32-1.98], *p*=0.001) and a 42% increased risk of all-cause mortality (HR 1.42 [1.26-1.60], *p*<0.0001) (**Table 4**). Each doubling of hs-cTnI was associated with a 53% increased risk of CVD mortality (HR 1.53 [1.35-1.72], *p*<0.0001) using the Abbott assay, a 44% increased risk using the Ortho assay (HR 1.44 [1.28-1.61], *p*<0.0001), and a 40% increased risk using the Siemens assay (HR 1.40 [1.26-1.55,] *p*<0.0001) (**Table 4**). Each doubling of NT-proBNP was associated with a 29% increased risk of CVD mortality (HR 1.29 [1.19-1.41], *p*<0.0001) and an 15% increased risk of all-cause mortality (HR 1.15 [1.09- 1.22], *p*<0.0001) (**Table 4**). Associations between cardiac biomarkers and non-CVD mortality were only significant for hs-cTnT and not hs-cTnI or NT-proBNP (**Table 4**). The linear relationship is demonstrated by restricted cubic splines in **Supplemental Figure 2**. There was no effect modification by CRP (*p-*interaction >0.05 for all cardiac biomarkers) and adding CRP to the models did not impact the effect estimates (data not shown).

**Table 4.** The association between eGFR and the difference between hs-cTnT and hs-cTnI.

|  | hs-cTnT – hs-cTnl<sub>Abbott</sub> |  | hs-cTnT – hs-cTnl<sub>Ortho</sub> |  | hs-cTnT – hs-cTnl<sub>Siemens</sub> |  |
| --- | --- | --- | --- | --- | --- | --- |
|  | Beta (95% CI) | p-value | Beta (95% CI) | p-value | Beta (95% CI) | p-value |
| **eGFR<sub>cr</sub>** |  |  |  |  |  |  |
| Model 1 | 7.8 (3.2, 12.5) | 0.002 | 8.5 (4.4, 12.7) | 0.0002 | 6.6 (-0.9, 14.1) | 0.08 |
| Model 2 | 7.2 (2.9, 11.5) | 0.002 | 7.7 (3.9, 11.5) | 0.0002 | 6.7 (-0.8, 14.2) | 0.08 |
| Model 3 | 6.6 (2.8, 10.5) | 0.001 | 7.1 (3.8, 10.4) | <0.0001 | 6.2 (-1.4, 13.7) | 0.11 |
| **eGFR<sub>cr-cys</sub>** |  |  |  |  |  |  |
| Model 1 | 8.5 (3.4, 13.6) | 0.002 | 9.1 (4.5, 13.7) | 0.0002 | 7.1 (-0.5, 14.7) | 0.07 |
| Model 2 | 7.7 (3.4, 11.9) | 0.0008 | 8.3 (4.4, 12.1) | <0.0001 | 7.0 (0.4, 13.6) | 0.04 |
| Model 3 | 7.8 (3.4, 12.1) | 0.0008 | 8.3 (4.4, 12.2) | <0.0001 | 7.1 (0.3, 13.8) | 0.04 |
| **eGFR<sub>cys</sub>** |  |  |  |  |  |  |
| Model 1 | 6.8 (3.4, 10.1) | 0.0002 | 7.5 (4.5, 10.5) | <0.0001 | 5.1 (-0.2, 10.3) | 0.06 |
| Model 2 | 6.1 (3.1, 9.2) | 0.0002 | 6.7 (3.9, 9.5) | <0.0001 | 5.0 (0.1, 9.9) | 0.04 |
| Model 3 | 6.2 (3.4, 9.0) | <0.0001 | 6.7 (4.2, 9.3) | <0.0001 | 5.1 (0.4, 9.9) | 0.04 |
| Model 1: Unadjusted.<br>Model 2: Model 1 + age, sex, race or ethnicity, smoking, health insurance, BMI, survey year, history of CVD, diabetes, hemoglobin A1c, total cholesterol, systolic blood pressure, ACEi/ARB use, statin use.<br>Model 3: Model 2 + UACR. |  |  |  |  |  |  |

**Table 5.**
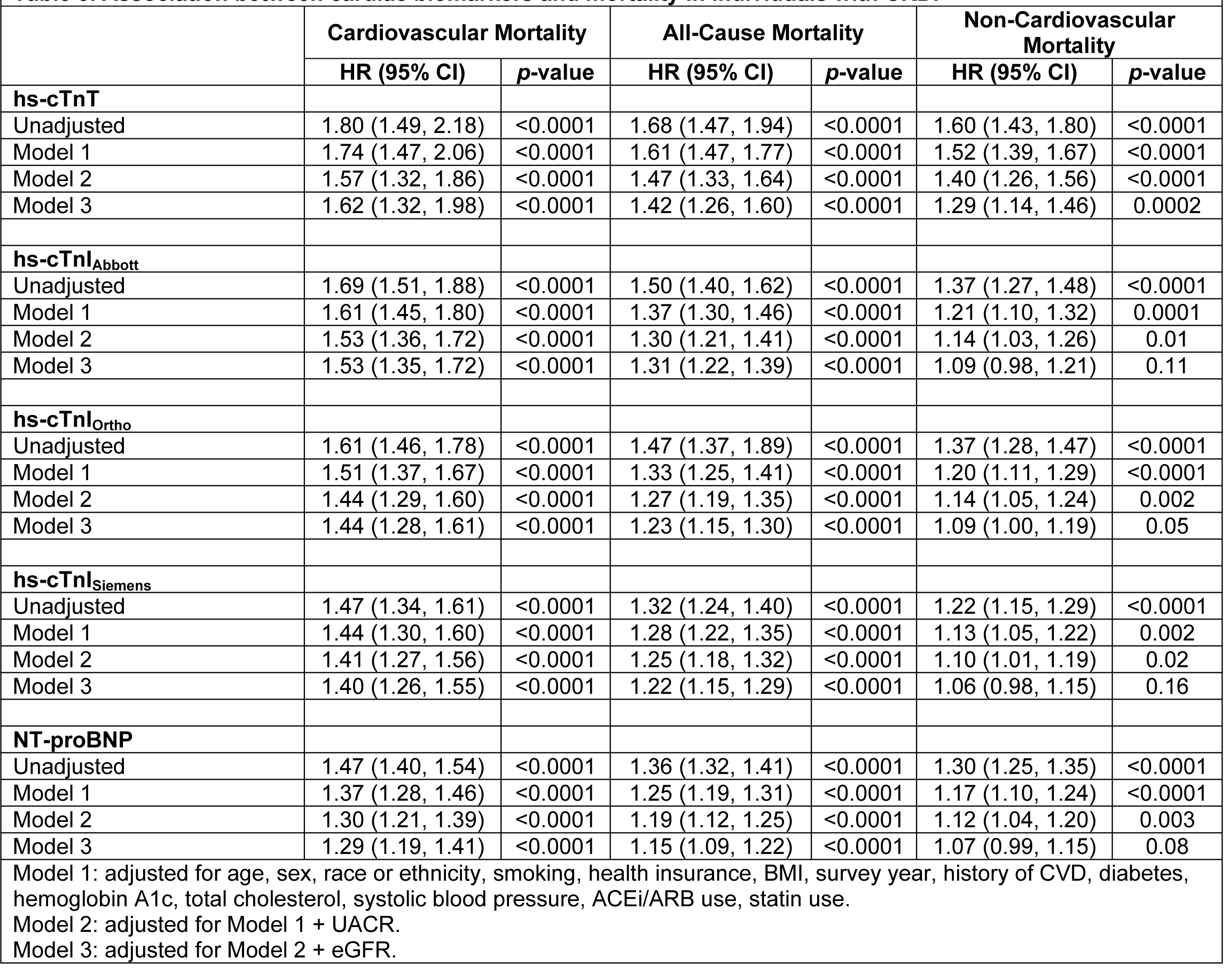
Association between cardiac biomarkers and mortality in individuals with CKD.

|  |  |  |  |  |  |  |
| --- | --- | --- | --- | --- | --- | --- |
|  | Cardiovascular Mortality |  | All-Cause Mortality |  | Non-Cardiovascular Mortality |  |
|  | HR (95% CI) | p-value | HR (95% CI) | p-value | HR (95% CI) | p-value |
| **hs-cTnT** |  |  |  |  |  |  |
| Unadjusted | 1.80 (1.49, 2.18) | <0.0001 | 1.68 (1.47, 1.94) | <0.0001 | 1.60 (1.43, 1.80) | <0.0001 |
| Model 1 | 1.74 (1.47, 2.06) | <0.0001 | 1.61 (1.47, 1.77) | <0.0001 | 1.52 (1.39, 1.67) | <0.0001 |
| Model 2 | 1.57 (1.32, 1.86) | <0.0001 | 1.47 (1.33, 1.64) | <0.0001 | 1.40 (1.26, 1.56) | <0.0001 |
| Model 3 | 1.62 (1.32, 1.98) | <0.0001 | 1.42 (1.26, 1.60) | <0.0001 | 1.29 (1.14, 1.46) | 0.0002 |
| **hs-cTnI<sub>Abbott</sub>** |  |  |  |  |  |  |
| Unadjusted | 1.69 (1.51, 1.88) | <0.0001 | 1.50 (1.40, 1.62) | <0.0001 | 1.37 (1.27, 1.48) | <0.0001 |
| Model 1 | 1.61 (1.45, 1.80) | <0.0001 | 1.37 (1.30, 1.46) | <0.0001 | 1.21 (1.10, 1.32) | 0.0001 |
| Model 2 | 1.53 (1.36, 1.72) | <0.0001 | 1.30 (1.21, 1.41) | <0.0001 | 1.14 (1.03, 1.26) | 0.01 |
| Model 3 | 1.53 (1.35, 1.72) | <0.0001 | 1.31 (1.22, 1.39) | <0.0001 | 1.09 (0.98, 1.21) | 0.11 |
| **hs-cTnI<sub>Ortho</sub>** |  |  |  |  |  |  |
| Unadjusted | 1.61 (1.46, 1.78) | <0.0001 | 1.47 (1.37, 1.89) | <0.0001 | 1.37 (1.28, 1.47) | <0.0001 |
| Model 1 | 1.51 (1.37, 1.67) | <0.0001 | 1.33 (1.25, 1.41) | <0.0001 | 1.20 (1.11, 1.29) | <0.0001 |
| Model 2 | 1.44 (1.29, 1.60) | <0.0001 | 1.27 (1.19, 1.35) | <0.0001 | 1.14 (1.05, 1.24) | 0.002 |
| Model 3 | 1.44 (1.28, 1.61) | <0.0001 | 1.23 (1.15, 1.30) | <0.0001 | 1.09 (1.00, 1.19) | 0.05 |
| **hs-cTnI<sub>Siemens</sub>** |  |  |  |  |  |  |
| Unadjusted | 1.47 (1.34, 1.61) | <0.0001 | 1.32 (1.24, 1.40) | <0.0001 | 1.22 (1.15, 1.29) | <0.0001 |
| Model 1 | 1.44 (1.30, 1.60) | <0.0001 | 1.28 (1.22, 1.35) | <0.0001 | 1.13 (1.05, 1.22) | 0.002 |
| Model 2 | 1.41 (1.27, 1.56) | <0.0001 | 1.25 (1.18, 1.32) | <0.0001 | 1.10 (1.01, 1.19) | 0.02 |
| Model 3 | 1.40 (1.26, 1.55) | <0.0001 | 1.22 (1.15, 1.29) | <0.0001 | 1.06 (0.98, 1.15) | 0.16 |
| **NT-proBNP** |  |  |  |  |  |  |
| Unadjusted | 1.47 (1.40, 1.54) | <0.0001 | 1.36 (1.32, 1.41) | <0.0001 | 1.30 (1.25, 1.35) | <0.0001 |
| Model 1 | 1.37 (1.28, 1.46) | <0.0001 | 1.25 (1.19, 1.31) | <0.0001 | 1.17 (1.10, 1.24) | <0.0001 |
| Model 2 | 1.30 (1.21, 1.39) | <0.0001 | 1.19 (1.12, 1.25) | <0.0001 | 1.12 (1.04, 1.20) | 0.003 |
| Model 3 | 1.29 (1.19, 1.41) | <0.0001 | 1.15 (1.09, 1.22) | <0.0001 | 1.07 (0.99, 1.15) | 0.08 |
| Model 1: adjusted for age, sex, race or ethnicity, smoking, health insurance, BMI, survey year, history of CVD, diabetes, hemoglobin A1c, total cholesterol, systolic blood pressure, ACEi/ARB use, statin use. |  |  |  |  |  |  |
| Model 2: adjusted for Model 1 + UACR. |  |  |  |  |  |  |
| Model 3: adjusted for Model 2 + eGFR. |  |  |  |  |  |  |

Adjusting for hs-cTnT in the models of hs-cTnI or NT-proBNP and mortality resulted in a slight reduction in the hazard ratios, but the associations remained statistically significant (**Supplemental Tables 2 and 3**). The c-statistics increased marginally when adjusting for hs-cTnT in models of CVD mortality (**Supplemental Table 2**). Stratifying by baseline history of CVD did not substantially impact the results and *p*-interaction was >0.05 (**Supplemental Table 4**).

## Discussion

In this nationally representative cohort of adults with CKD, hs-cTnI did not vary across eGFR as much as hs-cTnT and NT-proBNP, suggesting the hs-cTnI may be more stable across a lower range of eGFR within CKD (<60 ml/min per 1.73m^2^). Furthermore, all three biomarkers (hs-cTnT, hs-cTnI, and NT-proBNP) provided prognostic information for community-dwelling adults with CKD. This may indicate that biomarker elevations imply underlying pathology and do not solely reflect impaired clearance in the setting of kidney disease.

In our analysis, hs-cTnI was less variable across clinical eGFR categories in CKD, suggesting that it may be preferred for clinical use in patients with CKD over hs-cTnT. We demonstrated this in our analysis of the 99^th^ percentiles of each biomarker as well as regression models showing eGFR was less strongly associated with hs-cTnI than hs-cTnT. Specifically, hs- cTnI_Siemens_ was least associated with eGFR, regardless of estimating equation used. In contrast, hs-cTnT was strongly associated with eGFR. This was previously seen in a smaller study of Swedish adults, where hs-cTnT increased more prominently at lower measured GFR than hs-cTnI (n=489).^17^ These and our findings suggest that hs-cTnI is less impacted by eGFR and therefore may better suited for clinical use than hs-cTnT in CKD.

Of the three troponin subunits in the troponin complex, the T and I subunits are specific to cardiac myocyte damage and therefore used in clinical practice.^18^ Troponin I is degraded and fragmented in the serum into several smaller fragments and the half-life of these fragments can vary substantially.^18,19^ Several assays have been developed to detect differing epitopes of the troponin I subunit but have been infrequently compared in the literature.^14^ ^11^ Due to its smaller size, hs-cTnI may be more readily cleared by the kidney in patients with CKD than hs-cTnT. We observed higher differences between T and I at lower eGFR. We speculate that there may be differences in the size of T and I that may be reflected in the clearance.^18^ For example, the fragment of hs-cTnI detected by the Siemens assay may be more similar in size to hs-cTnT. However, additional work is needed to clarify these relationships as the exact size of the fragments identified by each assay and the mechanisms of kidney metabolism and filtration of these fragments are not well understood.

Cardiac biomarker concentrations were higher than the upper limit of normal in the general population and previously proposed upper reference limits in CKD.^20,21^ This is concordant with prior work demonstrating an association between eGFR and cardiac biomarker concentrations.^1,2,17,22^ However, it does not explain the mechanism behind this finding, which has previously been attributed to impaired renal clearance of these biomarkers. Our analyses of the difference between hs-cTnT and hs-cTnI concentration and eGFR suggest that the differences between the concentrations of these biomarkers is largest at the lowest eGFR. This might support the argument that there are differences in the clearance of hs-cTnT versus hs-cTnI.

Renal excretion is unlikely to be the sole mechanism responsible for elevated cardiac biomarker concentrations in CKD.^23–25^ Cardiac biomarker concentrations may be elevated due to an opposing process, such as increased production in the setting of chronic cardiac strain.^25^ Prior work has shown that increased hs-cTnT is cross-sectionally associated with increased UACR independent of eGFR.^26^ This finding may indicate that patients with worse CKD (e.g., higher UACR) have greater cardiac strain and therefore troponin leak. In children and adolescents with advanced CKD, elevated cardiac troponin T was attributed to ongoing myocardial injury rather than decreased clearance alone due to poor left-ventricular contractility.^27^ Smoldering, subclinical CVD and endothelial dysfunction may be present years prior to clinically evident changes in cardiac structure or function among patients with CKD.

Understanding the diagnostic, prognostic, and clinical implications of these assays is critical to clinical care. In patients with CKD and chronically elevated troponin concentrations in the absence of acute myocardial ischemia, the leading hypothesis is that elevations are due to impaired renal clearance. Therefore, the measured concentration is often overlooked in favor of examining a ‘delta’ or trend in the appropriate clinical setting.^17,22^ Our findings validate prior research showing that single time point elevations in biomarker concentrations are associated with increased risk of CVD mortality in CKD,^8,28,29^ suggesting that chronically elevated cardiac biomarkers in CKD may not be as benign as previously thought. Understanding the underlying pathology that leads to chronic biomarker elevations may yield important insights into CVD risk pathways in CKD. Diffuse endothelial damage may be driving the association with CVD mortality. Further work is needed to examine markers of endothelial dysfunction that are more mechanistically plausible than CRP, which is a generalized marker of inflammatory states.

Progressive CKD, accumulation of uremic toxins, and increased burden of CVD risk factors all increase the risk of CVD mortality and, independently, may increase systemic endothelial dysfunction.

There are several limitations to this analysis, including possible misclassification bias in administrative cause of death and reliance on one-time measurements of UACR and cardiac biomarkers. We were unable to adjust for interim CVD events in the mortality analyses as this information is not available in NHANES. The study also has several strengths, including the cohort demographic representation, inclusion of multiple biomarkers (including hs-cTnI) and multiple assays of hs-cTnI, and the comparison of creatinine- and cystatin c-based eGFR estimating equations. We used only samples that had not undergone a prior freeze-thaw cycle to minimize analytical factors that could influence the results. Furthermore, the models are robustly developed and incorporate sociodemographic and clinical characteristics not captured in prior reports.

## Conclusions

In this study, community dwelling adults with CKD had elevated levels of cardiac biomarkers suggestive of subclinical myocardial injury. Differences in the performance of specific assays in CKD have clinical relevance. In this study, hs-cTnI was least affected by eGFR and retained prognostic significance, suggesting that it may be the superior assay for clinical use in CKD.

## Disclosures

None

### Data Availability

The data used in this manuscript are publicly available at https://wwwn.cdc.gov/nchs/nhanes/default.aspx.

---

**Supplemental Figure 1.**
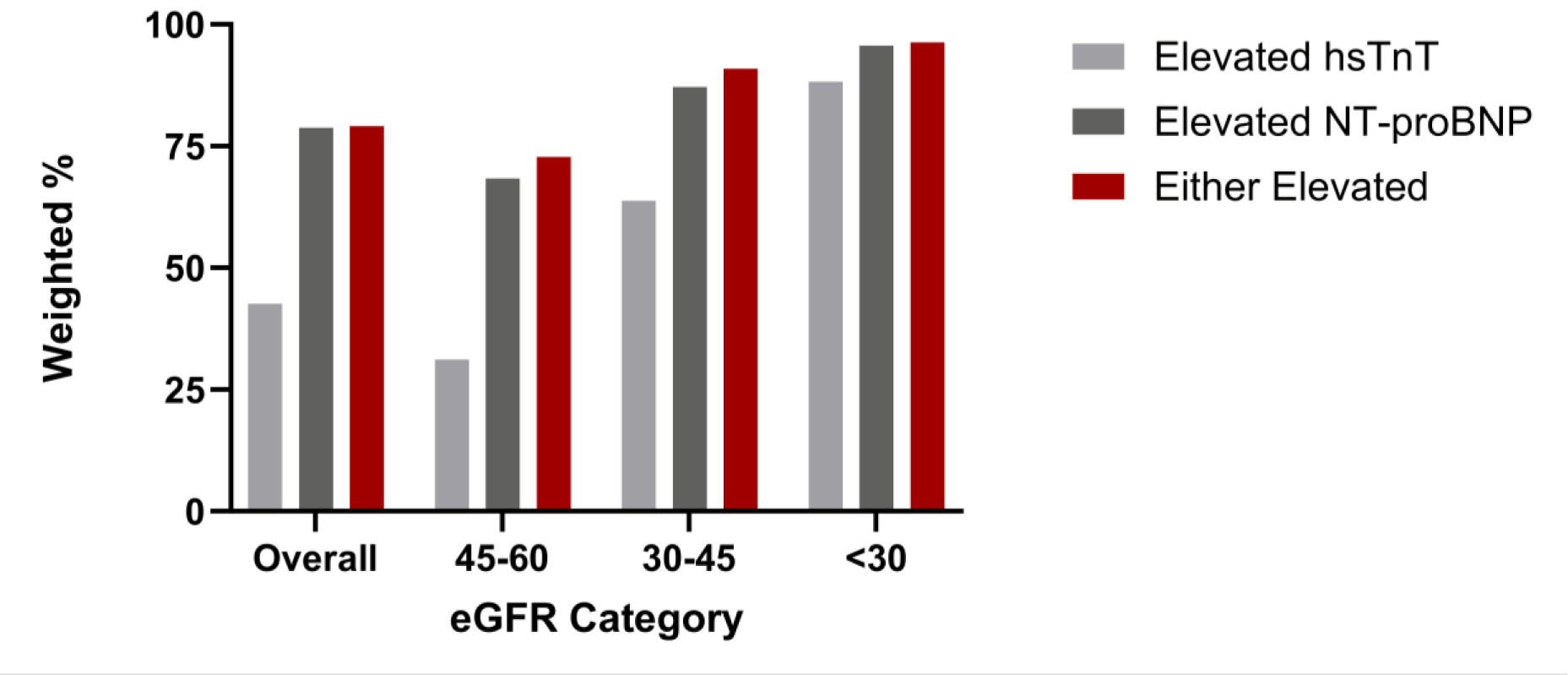
Prevalence of subclinical CVD by eGFR category in patients with CKD, using conventional thresholds from the general population. Thresholds include NT-proBNP ≥125 pg/mL and hs-cTnT ≥14 ng/L (women) or ≥22 ng/L (men).

**Supplemental Figure 2.**
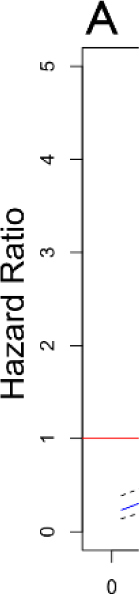
Restricted cubic splines of the relationship between cardiac biomarkers and cardiovascular disease mortality. A, hs-cTnT; B, hs-cTnI_Abbott_; C, NT-proBNP. Restricted cubic spline model adjusted for age, sex, race or ethnicity, smoking, health insurance, BMI, survey year, history of CVD, diabetes, hemoglobin A1c, total cholesterol, systolic blood pressure, ACEi/ARB use, statin use, UACR, eGFR.

**Supplemental Table 1.**
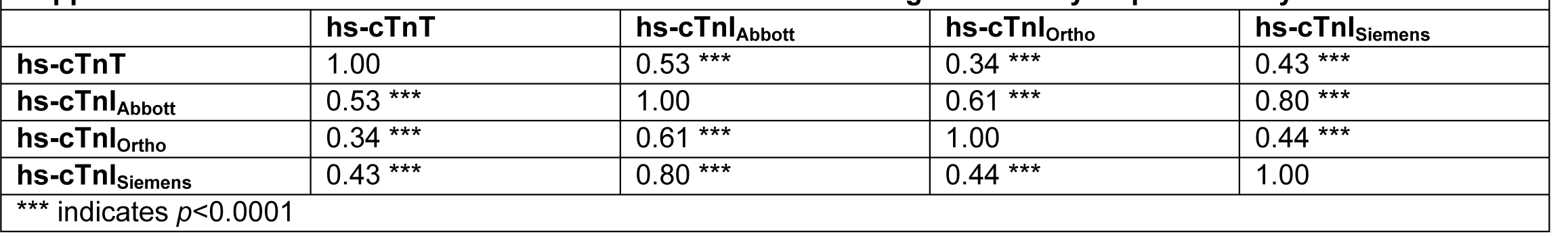
Pearson correlation coefficients between high sensitivity troponin assays.

**Supplemental Table 2.**
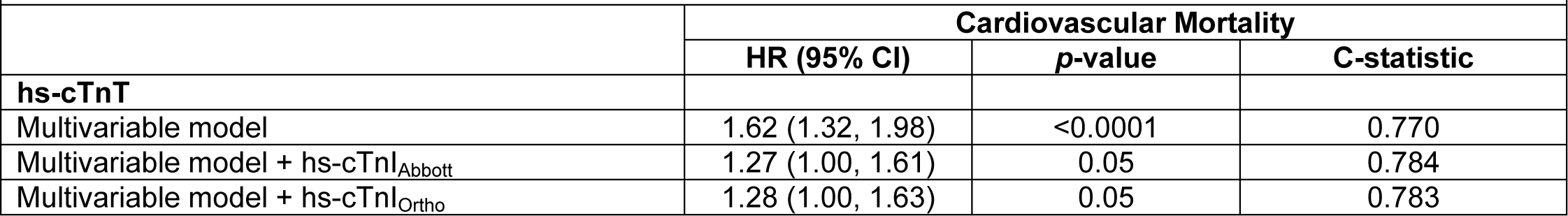
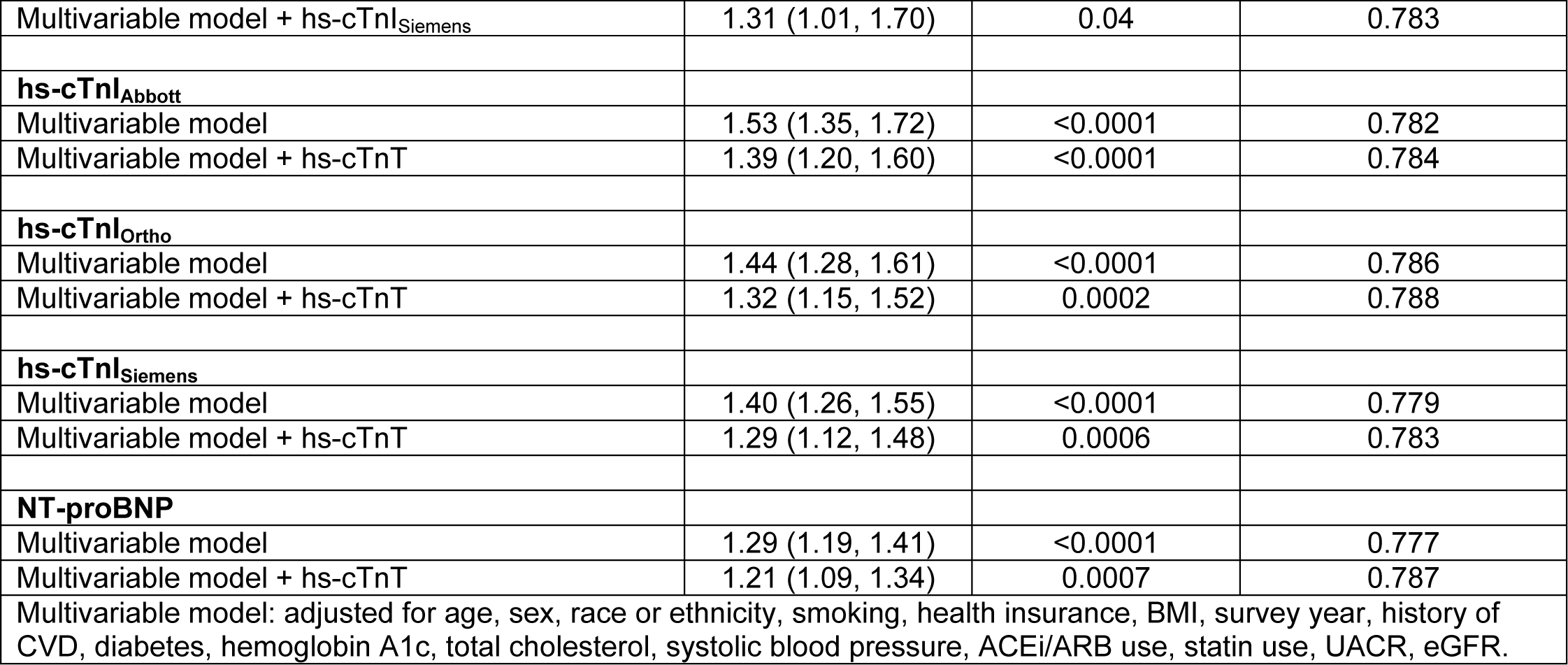
Association between cardiac biomarkers and cardiovascular mortality in individuals with CKD.

**Supplemental Table 3.** Association between cardiac biomarkers and non-cardiovascular and all-cause mortality in individuals with CKD.

|  |  |  |  |  |
| --- | --- | --- | --- | --- |
|  | **All-Cause Mortality** |  | **Non-Cardiovascular Mortality** |  |
|  | **HR (95% CI)** | **p-value** | **HR (95% CI)** | **p-value** |
| **hs-cTnT** |  |  |  |  |
| Multivariable model | 1.42 (1.26, 1.60) | <0.0001 | 1.29 (1.14, 1.46) | 0.0002 |
| Multivariable model + hs-cTnI (Abbott) | 1.29 (1.12, 1.49) | 0.0009 | 1.30 (1.09, 1.54) | 0.005 |
| Multivariable model + hs-cTnI (Ortho) | 1.29 (1.12, 1.48) | 0.0009 | 1.29 (1.09, 1.52) | 0.004 |
| Multivariable model + hs-cTnI (Siemens) | 1.29 (1.12, 1.50) | 0.0009 | 1.28 (1.10, 1.49) | 0.003 |
| **hs-cTnI<sub>Abbott</sub>** |  |  |  |  |
| Multivariable model | 1.31 (1.22, 1.39) | <0.0001 | 1.09 (0.98, 1.21) | 0.11 |
| Multivariable model + hs-cTnT | 1.15 (1.05, 1.26) | 0.004 | 0.98 (0.86, 1.13) | 0.81 |
| **hs-cTnI<sub>Ortho</sub>** |  |  |  |  |
| Multivariable model | 1.23 (1.15, 1.30) | <0.0001 | 1.09 (1.00, 1.19) | 0.05 |
| Multivariable model + hs-cTnT | 1.13 (1.03, 1.24) | 0.008 | 1.00 (0.89, 1.12) | 0.99 |
| **hs-cTnI<sub>Siemens</sub>** |  |  |  |  |
| Multivariable model | 1.22 (1.15, 1.29) | <0.0001 | 1.06 (0.98, 1.15) | 0.16 |
| Multivariable model + hs-cTnT | 1.11 (1.03, 1.21) | 0.008 | 0.99 (0.90, 1.09) | 0.81 |
| **NT-proBNP** |  |  |  |  |
| Multivariable model | 1.15 (1.09, 1.22) | <0.0001 | 1.07 (0.99, 1.15) | 0.08 |
| Multivariable model + hs-cTnT | 1.10 (1.02, 1.17) | 0.01 | 1.02 (0.95, 1.11) | 0.56 |
| Multivariable model: adjusted for age, sex, race or ethnicity, smoking, health insurance, BMI, survey year, history of CVD, diabetes, hemoglobin A1c, total cholesterol, systolic blood pressure, ACEi/ARB use, statin use, UACR, eGFR. |  |  |  |  |

**Supplemental Table 4.** Stratified association between cardiac biomarkers and mortality in individuals with CKD.

| Supplemental Table Stratified association between cardiac biomarkers and mortality in individuals with CKD. |  |  |  |  |  |  |
| --- | --- | --- | --- | --- | --- | --- |
|  | Cardiovascular Mortality |  |  | All-Cause Mortality |  |  |
|  | *p*-interaction | HR (95% CI) | *p*-value | *p*-interaction | HR (95% CI) | *p*-value |
| **hs-cTnT** | 0.23 |  |  | 0.25 |  |  |
| Prior CVD History |  | 1.59 (1.26, 2.02) | 0.0003 |  | 1.44 (1.24, 1.69) | <0.0001 |
| No Prior CVD History |  | 1.79 (1.33, 2.41) | 0.0003 |  | 1.47 (1.25, 1.73) | <0.0001 |
| **hs-cTnI<sub>Abbott</sub>** | 0.66 |  |  | 0.97 |  |  |
| Prior CVD History |  | 1.65 (1.39, 1.95) | <0.0001 |  | 1.31 (1.17, 1.45) | <0.0001 |
| No Prior CVD History |  | 1.43 (1.20, 1.69) | 0.0002 |  | 1.24 (1.11, 1.38) | 0.0003 |
| **hs-cTnI<sub>Ortho</sub>** | 0.72 |  |  | 0.73 |  |  |
| Prior CVD History |  | 1.51 (1.27, 1.79) | <0.0001 |  | 1.23 (1.11, 1.37) | 0.0002 |
| No Prior CVD History |  | 1.36 (1.16, 1.59) | 0.0003 |  | 1.22 (1.09, 1.36) | 0.0007 |
| **hs-cTnI<sub>Siemens</sub>** | 0.89 |  |  | 0.87 |  |  |
| Prior CVD History |  | 1.45 (1.25, 1.69) | <0.0001 |  | 1.22 (1.11, 1.33) | <0.0001 |
| No Prior CVD History |  | 1.33 (1.13, 1.57) | 0.001 |  | 1.18 (1.08, 1.28) | 0.0003 |
| **NT-proBNP** | 0.01 |  |  | 0.04 |  |  |
| Prior CVD History |  | 1.21 (1.10, 1.34) | 0.0002 |  | 1.13 (1.06, 1.21) | 0.0005 |
| No Prior CVD History |  | 1.40 (1.22, 1.60) | <0.0001 |  | 1.18 (1.08, 1.30) | 0.0008 |
| Models are adjusted for age, sex, race or ethnicity, smoking, health insurance, BMI, survey year, history of CVD, diabetes, hemoglobin A1c, total cholesterol, systolic blood pressure, ACEi/ARB use, statin use, UACR, and eGFR. |  |  |  |  |  |  |

